# COVID-19 RT-PCR diagnostic assay sensitivity and SARS-CoV-2 transmission: A missing link?

**DOI:** 10.1101/2021.03.24.21254271

**Authors:** Nikhil Shri Sahahjpal, Ephrem Chin Lip Hon, Stephanie Dallaire, Colin Williams, Sudha Ananth, Ashis K Mondal, Amyn M Rojiani, Madhuri Hegde, Ravindra Kolhe

## Abstract

**Background:** The sensitivity of commercially available RT-PCR assays varies over 10,000 fold, ranging from 10 to 20,000 viral copies/ml. The reporting of high Ct value results has been under scrutiny, as the clinical significance of these values is not yet completely understood. The early detection of infected individuals (high Ct results) in the pre-symptomatic phase of the disease using highly sensitive RT-PCR methods has been argued as a strategy to prevent transmission, while on the contrary, the reporting of high Ct has been criticized as false-positive results causing unnecessary testing and having several negative implications. The purpose of this study was to verify the presence of SARS-CoV-2 genomes in samples with a wide range of RT-PCR Ct values including samples with high Ct (37 to 42) using next-generation sequencing (NGS).

**Methods:** The study evaluated a total of 547 previously positive samples tested with the PerkinElmer® New Coronavirus Nucleic Acid Detection RT-PCR kit. The samples included in this study ranged from Ct values of 17-42, with 44 samples having a Ct > 37. Of the 547 samples, 149 were sequenced using PerkinElmer NEXTFLEX Variant-Seq SARS-CoV2 assay on NovaSeq 6000, and 398 samples were sequenced using Illumina SARS-CoV-2 respiratory viral panel kits using the NextSeq 500/550 system.

**Results:** Between the two clinical laboratories, a total of ∼1.95 million samples were tested using the FDA-EUA PerkinElmer® New Coronavirus RT-PCR assay. Of the 1.95 million samples, ∼1.72 million were negative, ∼250,000 positive, and ∼16,500 in the range of 37-42. Of the 547 samples sequenced, the percentage of sequencing reads that aligned to the SARS-CoV-2 Wuhan-hu-1 reference genome (NC_045512.2) ranged from 25.5% to 99.69%. All samples sequenced showed high sequence specificity to the SARS-CoV-2 virus. Low Ct samples showed complete uniform coverage across the entire 29kb SAR-CoV-2 genome. The average coverage in samples with high Ct (>37) was found to be 55.5% (range 16.1-99.2%). However, as sample Ct increased, a gradual decrease in coverage uniformity was observed for few samples.

**Conclusion:** This study demonstrates for the first time that the viral RNA is present in the high Ct value range of 37-42 and the sequence is unique to SARS-CoV-2 confirmed using two separate sequencing assays. This confirms that the detected Ct values are reflective of the presence of the SARS-CoV-2 virus and they are not an artifact or contamination. In light of the recent work highlighting the majority of transmission being pre-symptomatic/ asymptomatic, and high Ct results being observed at both the early and late phases of infection warrants further investigation into the clinical utility of high Ct results to curtail the spread of the virus.

## Introduction

The COVID-19 outbreak caused by SARS-CoV-2 is currently a rampant pandemic and has led to major socio-economic disruption worldwide. Since the identification of SARS-CoV-2 in the region of Wuhan, China, 122,941,299 confirmed cases with over 2,711,650 COVID-19 related deaths have been reported globally (https://coronavirus.jhu.edu/map.html, last accessed March 21, 2021), far exceeding the previous SARS-CoV epidemic, and influenza pandemics (2009 and 1918). Several viral and host characteristics such as high transmissibility (R_0_ 2.5), the incubation period (4-12 days), and a high proportion of asymptomatic/mild illness cases might have accounted for the spread of SARS-CoV-2 virus as compared to SARS-CoV and influenza viruses. ^1^ Testing for SARS-CoV-2 has been identified as the single most important measure to curtail the spread of the disease, as it is the primary step that enables the effectiveness of government strategies such as quarantine, contact tracing, travel restrictions, and lockdowns.^2^ Currently, RT-PCR-based assays are the predicate method for detecting SARS-CoV-2, targeting selected regions of the virus nucleocapsid (N), envelop (E), spike (S), and/or open reading frame (ORF) genes.^3^ The RT-PCR methodology is inherently quantitative but U.S. Food and Drug Administration (FDA) Emergency Use Authorized (EUA) RT-PCR tests for the detection of SARS-CoV-2 are designed as qualitative assays. The limit of detection (LOD) of the RT-PCR assays available commercially ranges from 8-10 viral copies/ml to 20,000 viral copies/ml that corresponds to approximately the cycle threshold (Ct) values of 45 to 10-11, respectively **(Figure 1)**.

**Figure 1.**
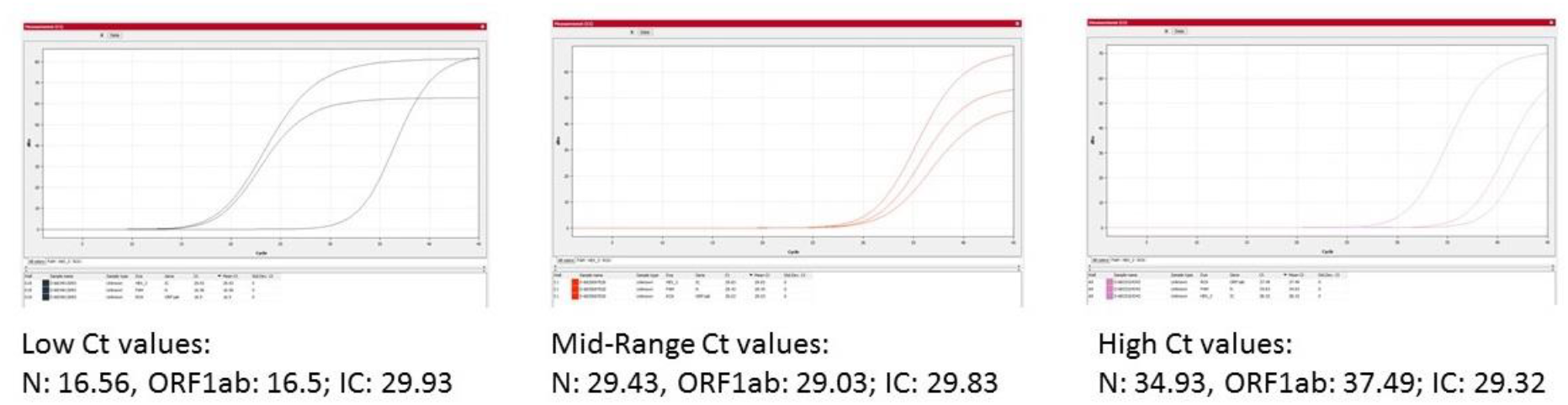
Example of RT-PCR traces of 3 positive RT-PCR samples with a low, mid-range and high Ct

Several groups have used RT-PCR Ct values to predict progression to severe COVID-19 disease and infer transmissibility. ^4-7^ Although these diagnostic assays are not normalized to controls with known concentration, an estimate of the viral load can be inferred using the Ct value, as the Ct value and viral load are inversely related. ^8^ Low Ct has been associated with a higher risk for severe COVID-19,^6,7^ whereas, high Ct has been associated with pre-as well as post-symptomatic phases of the disease.^9,10^ Appallingly, decision analytical models have assessed that 59% of all transmission are asymptomatic transmission, comprising 35% from pre-symptomatic and 24% from asymptomatic individuals. ^11^ However, measures to reduce transmission from individuals who do not have COVID-19 symptoms have been politicized as these likely had negative effects on the economy. Further, the reporting of high Ct ranges has been questioned and debated as the importance of these values with regards to viral load is not yet completely understood. The early detection of infected individuals (high Ct results) in the pre-symptomatic phase of the disease using highly sensitive RT-PCR methods has been argued as a strategy to prevent transmission,^10^ while on the contrary, the reporting of high Ct has been criticized as false-positive results causing unnecessary testing and having negative implications. ^12^

To date, the verification of the presence of the SARS-CoV-2 virus in the high Ct samples has not been demonstrated. It has been shown that the high Ct values >33 are not reproducible when an individual is re-tested within 24 hours and the reproducibility of the original Ct values is not consistent after 6 hours. It is necessary to evaluate the presence of the virus in these samples to do the clinical correlation and evaluate its association if any, to the spread and resurgence events we are experiencing globally. The purpose of this study was to verify the presence of SARS-CoV-2 genomes in samples with a wide range of RT-PCR cycle threshold (Ct) values including samples with high Ct (37 to 42) (recorded using RT-PCR method with LOD of 20 copies/ml) using next-generation sequencing (NGS).

## Materials and Methods

This multi-center diagnostic study was conducted at PerkinElmer Genomics (Site-1), CA, USA, and Augusta University (Site-2), GA, USA. The samples were processed under an approved HAC by the IRB Committee A (IRB REGISTRATION # 611298), Augusta University, GA. Based on the IRB approval, all PHI was removed and all data was anonymized before accessing for the study.

### Assay for the detection of SARS-CoV-2

The PerkinElmer® New Coronavirus Nucleic Acid Detection Kit is a real-time RT-PCR in vitro diagnostic test intended for the qualitative detection of nucleic acid from SARS-CoV-2 in human oropharyngeal swab and nasopharyngeal swab specimens collected by a healthcare provider (HCP), and anterior nasal swab specimens collected by an HCP or self-collected under the supervision of an HCP. The PerkinElmer® New Coronavirus Nucleic Acid Detection kit uses TaqMan-based real-time PCR technique to conduct in vitro reverse transcription of SARS-CoV-2 RNA, DNA amplification, and fluorescence detection. The assay targets specific genomic regions of the SARS-CoV-2: nucleocapsid (N) gene and Open Reading Frame 1ab (ORF1ab) gene. The TaqMan probes for the two amplicons are labeled with FAM and ROX fluorescent dyes respectively to generate a target-specific signal. The assay includes an RNA internal control (IC, bacteriophage MS2) to monitor the processes from nucleic acid extraction to fluorescence detection. The IC probe is labeled with VIC fluorescent dye to differentiate its fluorescent signal from SARS-CoV-2 targets. The assay also uses a dUTP/UNG carryover prevention system to avoid contamination of PCR products and subsequent false-positive results **(Figure 2**). The assay was validated by performing a limit of detection (LOD), inclusivity, cross-reactivity, and clinical evaluation studies.

**Figure 2.**
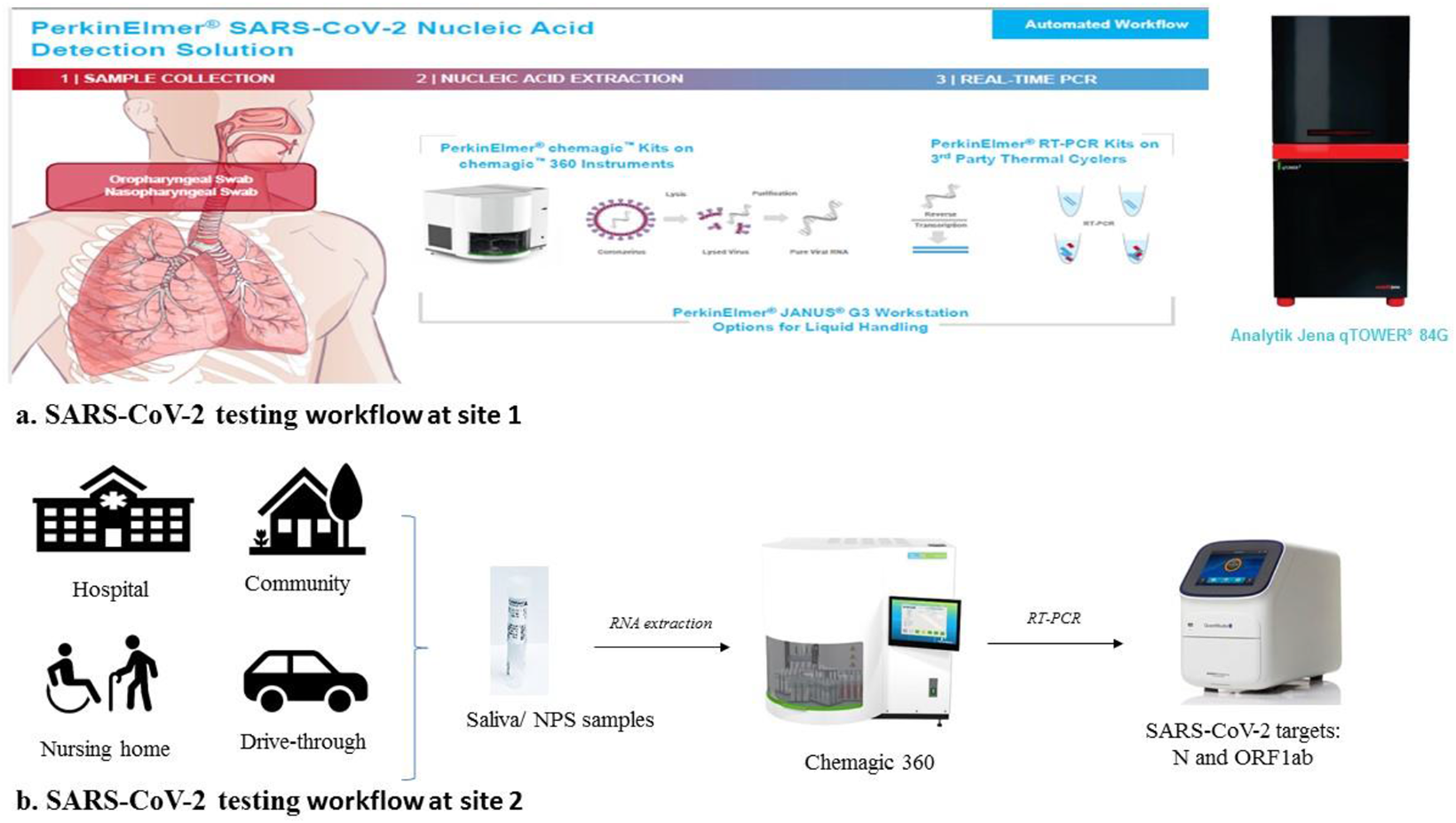
Figure 2 illustrates the simple RT-PCR workflow that has been established for COVID-19 testing. a. Site 1-Swab sample is extracted using an automated Chemagic360 robot that utilizes magnetic bead extraction technology. Extracted RNA sample is then combined with PerkinElmer® New Coronavirus Nucleic Acid Detection kit in a 384 well plate for real-time PCR and the detection of the SARS-CoV-2 virus using the Analytik Jena qTOWER system. b. NPS and/or saliva samples received from hospitals, communities, drive-throughs, and nursing homes are processed for RNA extraction using chemagic 360 and RT-PCR using Quantstudio 3 and 5 systems.

### Limit of Detection

LoD studies were defined for oropharyngeal and nasopharyngeal swab specimens using the PerkinElmer® Nucleic Acid Extraction kit on the PreNAT II Automated Workstation with The PerkinElmer® New Coronavirus Nucleic Acid Detection kit. This study was repeated using the chemagic Viral DNA/RNA 300 Kit special H96 (CMG-1033) on the chemagic 360 instruments. The limit of detection (LoD) studies was conducted as per the FDA guidelines (https://www.fda.gov/medical-devices/coronavirus-disease-2019-covid-19-emergency-use-authorizations-medical-devices/vitro-diagnostics-euas). The LoD for this RT-PCR kit was determined to be 20 copies/mL of SARS-CoV-2 in a sample.

### Inclusivity and Cross-reactivity of the PerkinElmer^®^ New Coronavirus Nucleic Acid Detection Kit

The inclusivity of the SARS-CoV-2 specific PerkinElmer ® New Coronavirus Nucleic Acid Detection kit primers/probes has been evaluated using full-length genomic sequences available in the NCBI and GISAID databases. Cross-reactivity of the PerkinElmer ^®^ New Coronavirus Nucleic Acid Detection Kit was evaluated using both *in silico* analysis and wet testing against normal and pathogenic organisms found in the respiratory tract. BLASTn analysis queries of the PerkinElmer^®^ New Coronavirus Nucleic Acid Detection Kit primers and probes were performed against public domain nucleotide sequences with default settings. The database search parameters were as follows: the match and mismatch scores were 1 and −3, respectively; the penalty to create and extend a gap in an alignment was 5 and 2, respectively, and the search parameters automatically adjusted for short input sequences and the expected threshold was 1000. The potential interference of the substances listed below was tested in both the presence and absence of SARS-CoV-2 RNA with the PerkinElmer^®^ New Coronavirus Nucleic Acid Detection Kit. SARS-CoV-2 positive samples were prepared by mixing each of the potentially interfering substances with the assay positive control (synthetic SARS-CoV-2 ORF1ab and N RNA template encapsulated in MS2 bacteriophage) at approximately 3× the LoD.

### Clinical Evaluation

The performance of the PerkinElmer® New Coronavirus Nucleic Acid Detection Kit was evaluated using contrived clinical oropharyngeal swabs and nasopharyngeal swabs. In addition to testing contrived specimens in the clinical matrix, the PerkinElmer® New Coronavirus Nucleic Acid Detection Kit was also evaluated using nasopharyngeal swab specimens collected from individuals suspected of COVID-19 infection. The clinical study included 32 positive and 30 negative NP samples, collected in VTM. For this study, 300 µL of each sample was extracted using the Chemagic 360 with the Chemagic Viral DNA/RNA 300 H96 Kit, eluting with 60µL elution buffer. A 40µL aliquot of the eluted nucleic acid was then used as input for the PCR reactions on the ABI 7500 standard PCR instrument as described in the instructions for use. Each of the patient samples was also tested with a highly sensitive RT-PCR comparator method. This demonstrates the analytical sensitivity and specificity of the assay.

### SARS-CoV-2 Sequencing

The study evaluated a total of 547 previously positive samples (stored aliquots) tested with the PerkinElmer® New Coronavirus Nucleic Acid Detection kit. The samples included in this study ranged from Ct values of 17-42, with 44 samples having a Ct > 37. Of the 547 samples, 149 were sequenced using PerkinElmer NEXTFLEX Variant-Seq SARS-CoV2 assay on NovaSeq 6000, and 398 samples were sequenced using Illumina SARS-CoV-2 respiratory viral panel kits using the NextSeq 500/550 system.

### PerkinElmer Sequencing assay

Briefly, eight microliters of undiluted extracted RNA was reverse transcribed and amplified using a publicly available SARS-CoV-2 genome-specific ARTIC v3 primer set (3) along with all reagents needed for reverse transcription, amplicon-based enrichment, and NGS library preparation using Unique Dual Index Barcodes (UDI, PerkinElmer). The resulting amplicon material (50-100 ng) was used to make NGS libraries using the NEXTFLEX® Rapid XP DNA-Seq kit and NEXTFLEX® RNA-Seq 2.0 UDI. Each library was then normalized to 10nM and equal volumes combined to make the final library pool. The final library pool was then prepared and loaded on to an Illumina® NovaSeq® 6000, per manufacturer’s instructions, using an Illumina® NovaSeq® 6000 SP Reagent Kit v1.5 (200 cycles) and flow cell and performing a paired-end run (2×75 bp) targeting ∼7M reads per library **(Figure 3)**

**Figure 3.**
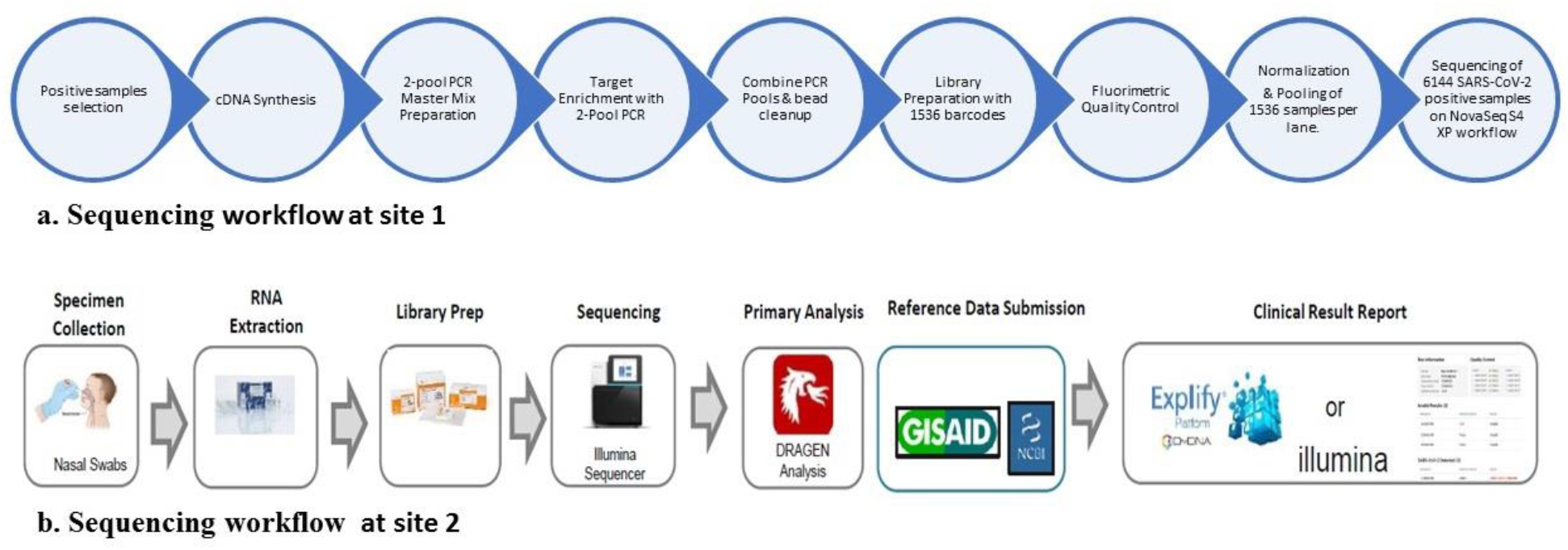
The figure illustrates the steps in the NGS workflow to generate libraries for sequencing. Site 1- With the availability of 1536 unique dual index and only needing to generate ∼1 million reads per sample, it is possible to run 6144 SARS-CoV-2 samples on the Illumina NovaSeq S4 flowcell. Site 2-With four index sets, A, B, C, and D each containing 96 unique, single-use Nextera UD indexes, using a 150bp paired-end sequencing approach and 300 cycles, the libraries were sequenced on the NextSeq 500/550 high-throughput sequencer using a V2 flowcell.

### Bioinformatics analysis

Data output in the form of FASTQ files were trimmed of adapters via Trimmomatic and aligned to the SARS-CoV-2 Wuhan-hu-1 reference genome (NC_045512.2) via Bowtie2 for bioinformatics analyses. To investigate cross-reactivity with other species alignment comparative analysis was performed by aligning sequences to 29 other bacterial and viral species. Alignment rates were plotted in a heatmap using Python scripting to visualize any potential signaling with species other than SARS-CoV-2.

### Illumina SARS-CoV-2 respiratory viral panel sequencing

Library preparation was performed following the Illumina RNA Prep, Tagmentation with enrichment protocol, and Nextera RNA kits. Briefly, 8.5 uL of extracted RNA, by the methodology described above, was denatured followed by first and second-strand DNA synthesis. This was followed by tagmentation, which uses enrichment bead-linked transposomes to tagment double-stranded cDNA. This process fragments cDNA and adds adapter sequences. After tagmentation, the fragments were purified and amplified to add index adapter sequences for dual indexing, and P7 and P5 sequences for clustering. Four index sets, A, B, C, and D each containing 96 unique, single-use Nextera UD indexes were used. Following clean-up, libraries were quantified using Invitrogen Qubit dsDNA broad range Assay Kit (Thermo Fischer Scientific). Subsequently, 7.5ul of the library was used for hybridization using oligos from the respiratory viral panel. This was followed by bead-based capture of hybridized probes, amplification, clean-up, and quantification of the enriched library. Normalized libraries diluted to an equimolar concentration of 0.8pM were then pooled into three runs. Using a 150bp paired-end sequencing approach and 300 cycles, the libraries were sequenced on the NextSeq 500/550 high-throughput sequencer using a V2 flow cell kit (Illumina).

### Bioinformatics analysis

Run metrics were evaluated on the Basespace app by analyzing cluster density and Q30 score. Sequences were then submitted for analysis to the Dragen pipeline for pathogen detection, coverage, and alignment to the SARS-CoV-2 genome available on the Basespace app.

## Results

A total of ∼1.95 million samples were tested at both sites (PerkinElmer and Augusta University, USA) using the PerkinElmer® New Coronavirus Nucleic Acid Detection kit. Of the 1.95 million samples, ∼1.72 million were negative, ∼250,000 positive, and ∼16,500 in the range of 37-42.

### PerkinElmer Sequencing assay

A total of 149 RT-PCR positive samples with Ct values spanning 17 to 42 were sequenced. This includes 38 samples with a Ct value of more than 37. The percentage of sequencing reads that aligned to the SARS-CoV-2 Wuhan-hu-1 reference genome (NC_045512.2) ranged from 38.69% to 99.69%. All samples sequenced showed high sequence specificity to the SARS-CoV-2 virus. No sample had 0% alignment to SARS-CoV-2 and no sample aligned better to any other species other than SARS-CoV-2. The average number of reads across the SARS-CoV-2 genome ranged from 275 to 1198 (average depth) with >90% of the 98 amplicons of the ARTIC v3 primer set represented. The bioinformatics analysis of NGS data for Ct values ranging from 17 to 42 for N and ORF1ab genes is shown in Table 1 **(Table 1)**.

**Table 1.**
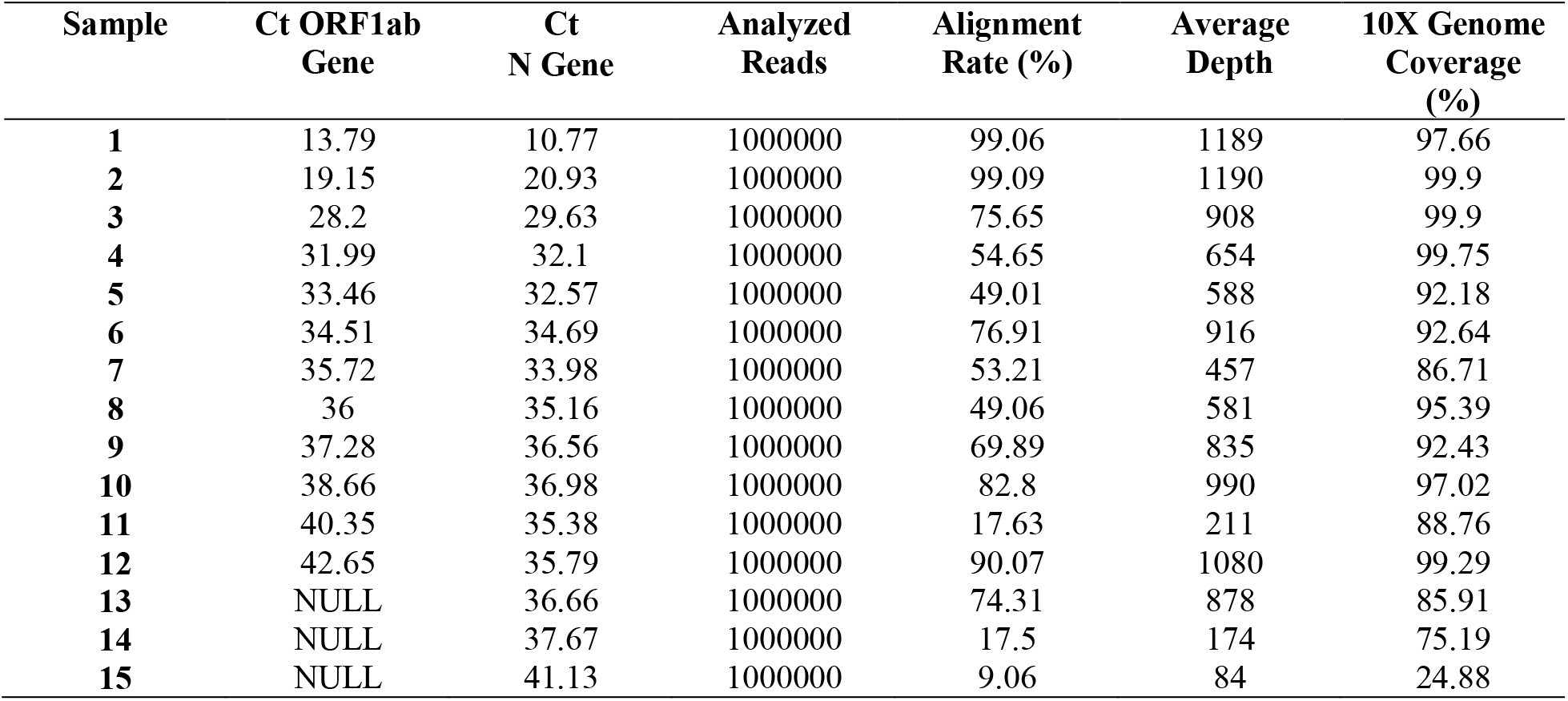
Representative of NGS performance data of RTPCR positive samples with ∼1M reads per sample. Samples span Ct values from 10 to 42 obtained using the PerkinElmer® New Coronavirus Nucleic Acid Detection kit. Sample 15 was the weakest of the entire sample set being sequenced but still obtained ∼25% SARS-CoV-2 genome coverage.

| Sample | Ct ORF1ab Gene | Ct N Gene | Analyzed Reads | Alignment Rate (%) | Average Depth | 10X Genome Coverage (%) |
| --- | --- | --- | --- | --- | --- | --- |
| 1 | 13.79 | 10.77 | 1000000 | 99.06 | 1189 | 97.66 |
| 2 | 19.15 | 20.93 | 1000000 | 99.09 | 1190 | 99.9 |
| 3 | 28.2 | 29.63 | 1000000 | 75.65 | 908 | 99.9 |
| 4 | 31.99 | 32.1 | 1000000 | 54.65 | 654 | 99.75 |
| 5 | 33.46 | 32.57 | 1000000 | 49.01 | 588 | 92.18 |
| 6 | 34.51 | 34.69 | 1000000 | 76.91 | 916 | 92.64 |
| 7 | 35.72 | 33.98 | 1000000 | 53.21 | 457 | 86.71 |
| 8 | 36 | 35.16 | 1000000 | 49.06 | 581 | 95.39 |
| 9 | 37.28 | 36.56 | 1000000 | 69.89 | 835 | 92.43 |
| 10 | 38.66 | 36.98 | 1000000 | 82.8 | 990 | 97.02 |
| 11 | 40.35 | 35.38 | 1000000 | 17.63 | 211 | 88.76 |
| 12 | 42.65 | 35.79 | 1000000 | 90.07 | 1080 | 99.29 |
| 13 | NULL | 36.66 | 1000000 | 74.31 | 878 | 85.91 |
| 14 | NULL | 37.67 | 1000000 | 17.5 | 174 | 75.19 |
| 15 | NULL | 41.13 | 1000000 | 9.06 | 84 | 24.88 |

Sequence information was obtained for all samples sequenced. Raw clusters per sample ranged from 365,690 up to 2,509,712. Samples with more than 1 million clusters were down-sampled for analysis. Low Ct samples showed complete uniform coverage across the entire 29kb SAR-CoV-2 genome. The average coverage in 38 samples with high Ct (>37) was found to be 55.5% (range 16.1-99.2%). However, as sample Ct increased, a gradual decrease in coverage uniformity was observed (**Figure 4**).

**Figure 4.**
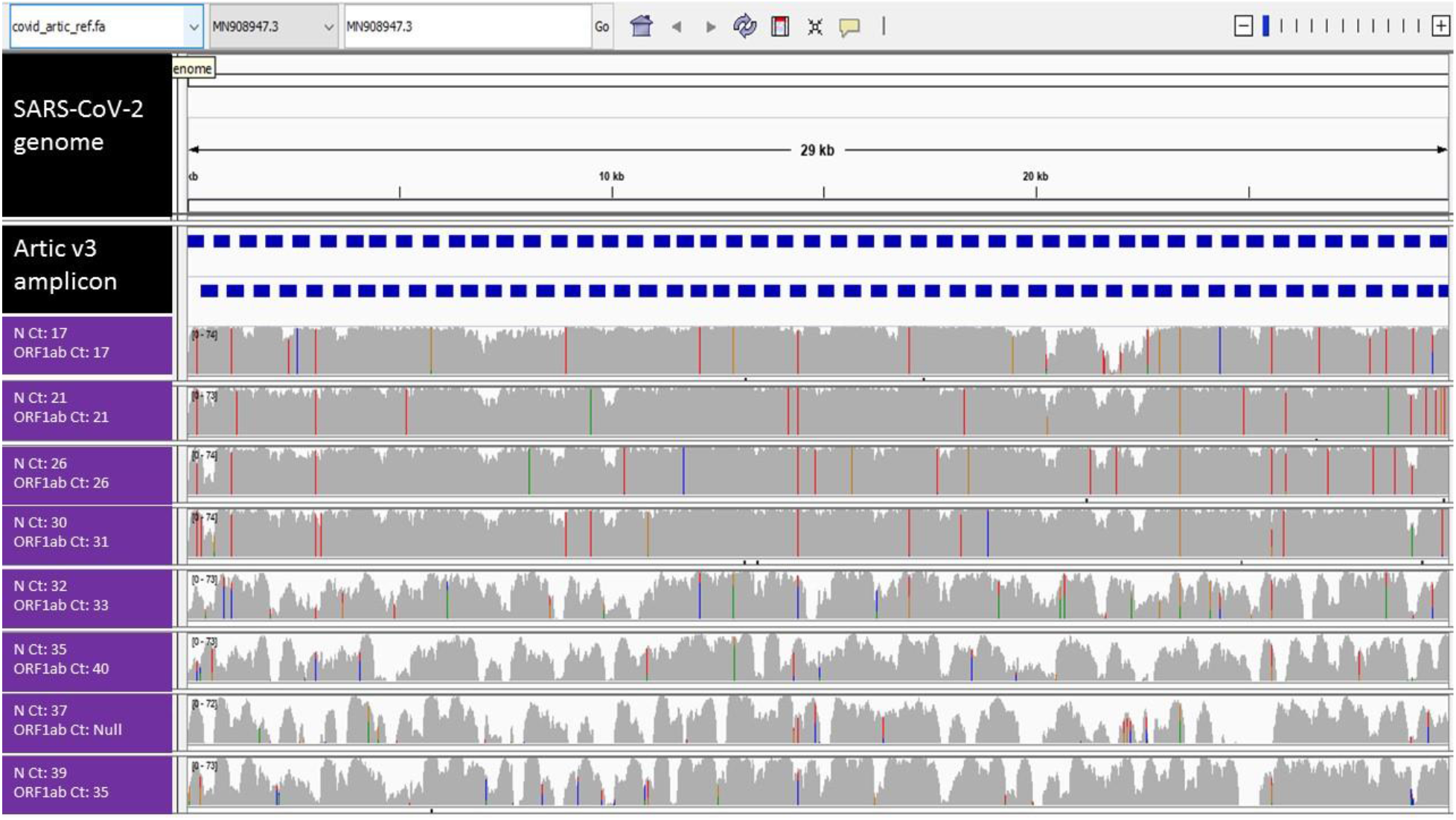
Representative coverage across the 29 kb SARS-CoV-2 genome of samples tested positive on PerkinElmer® New Coronavirus Nucleic Acid Detection kit. The top panel represents the SARS-CoV-2 WUHAN genomic sequence. The ARTIC V3 amplicons are presented in blue with pool A primers at the top and pool B primers below. The sequence alignment of the Ct values in N and ORF1b genes in the range of 17 to 40 are shown in the purple panels. The Ct value of 17 has the highest viral load (and possible most intact genomic content) which is demonstrated by the high depth of coverage @ ∼75X. Similar coverage was observed for Ct 30. Even though the bottom four panels (samples with Ct 32 to 40) did not have full genome sequence coverage, but there was unique SARS-CoV-2 genome data present indicating that there is SARS-CoV-2 virus present in the sample.

### Illumina SARS-CoV-2 respiratory viral panel sequencing

A total of 398 RT-PCR positive samples with Ct values spanning 10 to 42 were sequenced. This includes six samples with a Ct value of more than 37. The percentage of sequencing reads that aligned to the SARS-CoV-2 Wuhan-hu-1 reference genome (NC_045512.2) ranged from 25.5% to 99.8%. All samples sequenced showed high sequence specificity to the SARS-CoV-2 virus. No sample had 0% alignment to SARS-CoV-2 and no sample aligned better to any other species other than SARS-CoV-2 (**Table 2**).

**Table 2.** Representative of NGS performance data of RTPCR positive samples Illumina SARS-CoV-2 respiratory viral panel. Sample 9 was amongst the weakest of the entire sample set being sequenced but still obtained ∼ 90% SARS-CoV-2 genome coverage.

| Sample | Ct ORF1ab<br>Gene | Ct<br>N Gene | Alignment<br>Rate (%) |
| --- | --- | --- | --- |
| 1 | 15.59 | 16.42 | 99.94 |
| 2 | 19.50 | 20.73 | 99.83 |
| 3 | 29.34 | 26.69 | 99.74 |
| 4 | 29.29 | 30.42 | 80.61 |
| 5 | 38.098 | NULL | 99.82 |
| 6 | NULL | 36.89 | 99.80 |
| 7 | 37.8 | NULL | 63.31 |
| 8 | 34.316 | 33.04 | 97.01 |
| 9 | 39.678 | NULL | 90.47 |

### Comparative analysis with other genomes

No sample had 0% alignment to SARS-CoV-2 and no sample aligned better to any other species other than SARS-CoV-2 or hg38. There is a small amount of alignment to the SARS genome, which is due to sequence homology between the SARS-CoV-2 and SARS genomes. This heat map comparing the Sequencing data to other genomes (29 bacterial and viral genomes) demonstrates that the sequence obtained is unique to SARS-CoV-2 **(Figure 5)**. The bioinformatics analysis of NGS data with reference to the Wuhan sequence and the comparative analysis with other genomes confirmed the presence of SARS-CoV-2 in samples associated with a wide range of Ct values including high Ct values generated using the PerkinElmer® New Coronavirus Nucleic Acid Detection kit in the range of Ct >37→<42.

**Figure 5.**
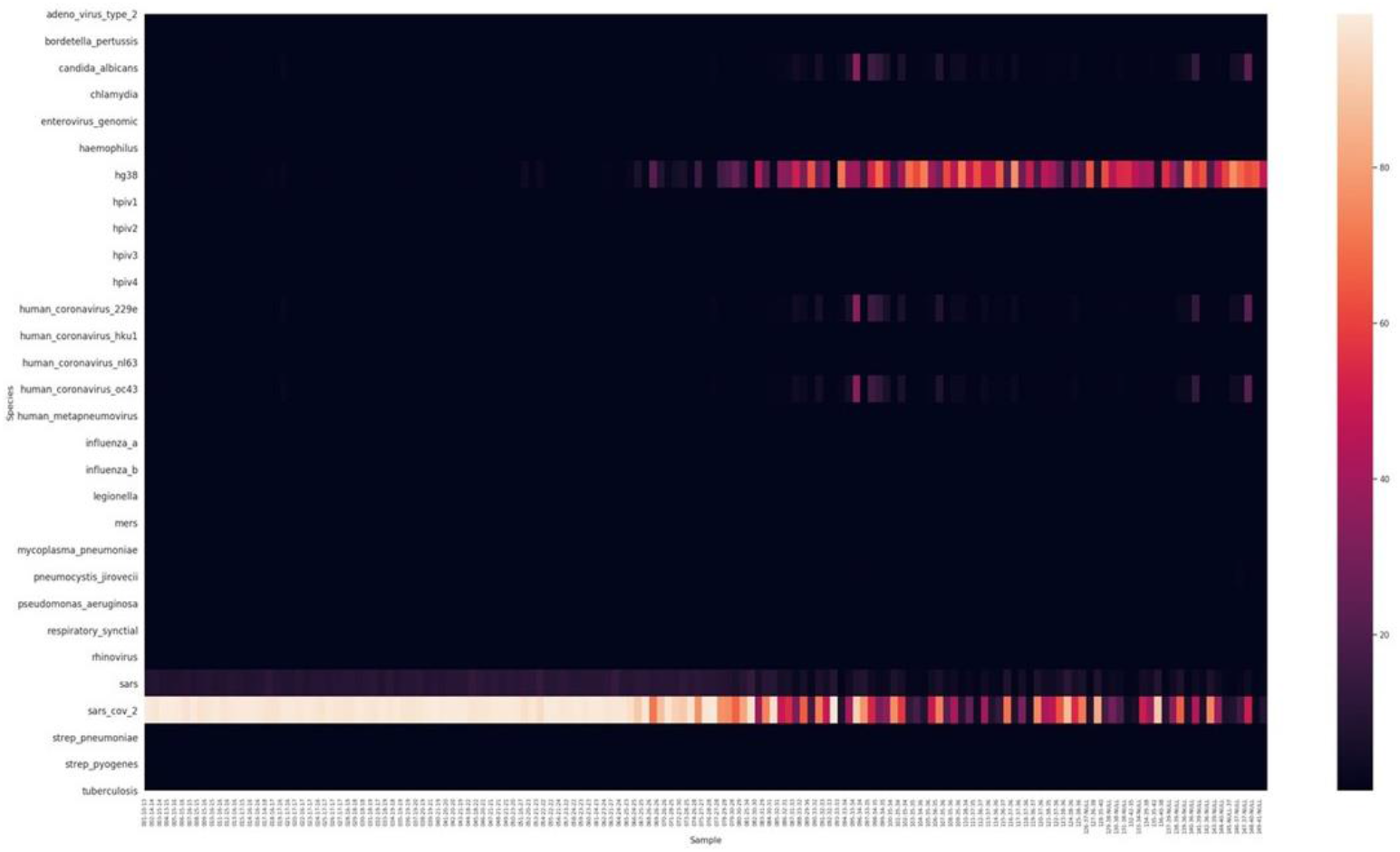
Alignment of reads, against 30 different species, generated from sequencing data of SAR-CoV-2 positive RNA samples demonstrating the unique sequence detection using the PerkinElmer New Coronavirus assay in low to high Ct ranges. Rows represent species, and columns represent samples sorted from low Ct to high Ct (left to right). The heatmap scale is from 0 to 100, representing the percent of sample reads aligning to each species. The dark area of the heat map indicates no alignment to the species listed on the left except for the SARS-CoV-2 and human hg38 genomes represented in patterned bars. Residual coverage, presumably artifacts or homologous sequences, were observed in candida albicans and two other human coronavirus species hku1 and oc43.

## Discussion

The World Health Organization (WHO) recognized COVID-19 as a global health emergency in late January 2021, which triggered manufacturer’s around the globe to develop molecular diagnostic assays for the detection of SARS-CoV-2 in clinical specimens. Since then, the RT-PCR-based assays have been the primary diagnostic tools, and numerous reports have compared the different assays for analytical sensitivity and specificity while addressing the complex factors that include sample collection, transport media, transport condition, and RNA extraction methods affecting the performance of these assay. Several other methods that include LAMP, End Point PCR, and ddPCR have also been described but are used in a limited capacity by some laboratories. The commercially available kits undergo regulatory evaluation such as CE-IVD declaration in Europe and FDA Emergency Use Authorization (EUA) in the US. To date, numerous assays have received EUA and CE-IVD and their sensitivity described as LOD ranges from 8-10 viral copies/ ml to 20,000 viral copies/ml (www.fda.gov). The FDA reference panel study conducted by the US FDA to compare EUA kits using the same reference control material demonstrated that the PerkinElmer® New Coronavirus Nucleic Acid Detection kit has the highest sensitivity of 180 NDU/ml. The next closest commercially available kit is 450 NDU/ml. This data reported on the FDA website shows that there is a 1000-fold difference in Limits of Detection (LOD) between theseassays.^13^ This has triggered an intense debate in the scientific community on the validity of the high Ct values and low viral load and the role they may or will play in the pandemic.

Several publications have demonstrated that about 3-25% of the high Ct results are not reproducible depending on the kit/assay used for repeat testing, with high Ct samples claimed as false positive with no clinical significance. ^14,15^ However, a significant caveat in extrapolating these studies to all RT-PCR methods is that these studies have not been able to take into account the variability in specimen collection techniques, collection types, and sample types used for collection and transport of the samples. Further, repeat studies on high Ct samples, which have undergone a freeze-thaw cycle, are likely to affect the RT-PCR assay sensitivity due to sample degradation. It is nearly impossible to compare the different kits/ assays due to their unique formulation, LoD claims, and the number of targets in the assay (1-4 targets that include combinations of N, E, ORF1ab, and S genes).

Multiple publications have demonstrated the correlation of the severity of COVID-19 disease to low Ct values and high viral loads, ^4,6,7^ whereas, the clinical utility of high Ct and low viral load is still being debated in the literature. Conflicting reports have emerged in the literature on the utility of high Ct results, with some reports demonstrating high Ct results as false positives, while others arguing the need to report high Ct results to prevent transmission. Recent data have shown that at these high Ct values retesting after 24 hours will very likely yield a negative result.^16^ Similar studies performed on samples with Ct >30 about 3- 25% of the samples retested as negative depending on the kit used.^17-19^ However, a recent study by Arnaut et al., highlights that the sensitivity of currently approved assay varies over 10,000-fold and several low sensitivity (high LoD) assays will miss the majority of infected patients that could be easily identified by assays having high sensitivity (low LoD). ^17^ Further, a study by Winnet et al., identifies a case of pre-symptomatic household transmission from a healthy young adult to a sibling and a parent, with high Ct (low viral load) results in the early phase (pre-symptomatic) of the infection that could prevent transmission in the family.^10^ Furthermore, an analysis of serial RT-PCR assays and chest CT scans has demonstrated that the mean interval between the initial negative to positive RT-PCR results was 5.1 days ± 1.5, and the mean interval between initial positive to subsequent negative RT-PCR results was 6.9 days ± 2.3.^20^ Thus, highly sensitive RT-PCR methods might be able to detect individuals in the early pre-symptomatic phase and prevent transmission in the community.

This study demonstrates for the first time that the viral RNA is present in the high Ct value range of ≥37 to <42 and the sequence is unique to SARS-CoV-2 from two independent laboratories using the PerkinElmer® New Coronavirus Nucleic Acid Detection Kit and two separate sequencing assays. This confirms that the detected Ct values are reflective of the presence of the SARS-CoV-2 virus and they are not an artifact or contamination. As shown in Fig. 4, the coverage of 29kb of the viral genome sequence is uniform at the lower Ct values for both the N and ORF1ab genes. The uniformity of coverage was observed to decreases for few samples as the Ct value increased. The aligned sequence of the high Ct values is indeed the conserved part of the SARS-CoV-2 genomic sequence. The depth and coverage as shown in tables 1 and 2, ranging from 16.1-99.2% and 100% to 10-15% which correlated as expected with low to high Ct values. We have also been able to demonstrate that almost 100% sequence obtained is unique to SARS-CoV-2 with minimal alignment to other human coronavirus species hku1 and oc43 and the candida albicans (Figure 5). Notably, this unique alignment was obtained despite the reduced coverage uniformity in the high Ct value samples.

It is importantto consider that if retesting is performed on a different assay, which has a LOD higher than the PerkinElmer® New Coronavirus Detection kit, the results are very likely to be negative. To date, despite extensive efforts, it has been difficult to control the increased number of COVID-19 cases. The question of the clinical utility of high Ct values in the context of COVID-19 remains unresolved, but it is clear from this work that the viral sequence is detectable in these samples, which demonstrates the analytical sensitivity and specificity of the assay. In light of the recent work highlighting majority (∼59%) of transmission being pre-symptomatic/asymptomatic, and high Ct results being observed at both the early and late phase of infection warrants further investigation into the clinical utility of high Ct results to curtail the spread of the virus.

## Data Availability

All relevant data has been included in the manuscript.

